# Evaluating Swiss growth reference curves: A comparative analysis in Zurich schoolchildren

**DOI:** 10.1101/2024.07.01.24309377

**Authors:** Lorenz M. Leuenberger, Fabiën N. Belle, Rebeca Mozun, Ben D. Spycher, Maria C. Mallet, Oskar G. Jenni, Christoph Saner, Philipp Latzin, Alexander Moeller, Claudia E. Kuehni, the LUIS Study Group

## Abstract

**INTRODUCTION:** Switzerland has an ongoing debate about the appropriateness of national growth reference curves. The Swiss Society of Pediatrics currently recommends the growth references of the World Health Organization (WHO), while the Center for Pediatric Endocrinology Zurich has proposed alternative growth references based on local data. Specialists and researchers also use International Obesity Task Force (IOTF) references to define overweight and obesity. We investigated the fit of these three growth references to anthropometric measurements from schoolchildren in the canton of Zurich and assessed the prevalence of overweight, obesity, and short stature across the three references.

**METHODS:** We analyzed data from 3755 children aged 6-17 years of the cross sectional LuftiBus in the school (LUIS) study, collected between 2013-2016 in the canton of Zurich. We calculated z-scores of height, weight, and body mass index (BMI) based on WHO, local, and IOTF references. We compared the mean and distribution of z-scores to the expected standard normal distribution using the Anderson-Darling test. We classified BMI based on cutoff values given by the three references: overweight (WHO: >90.0^th^ percentile; local: >82.9_[girls]_, >78.9_[boys]_; IOTF: >89.3_[girls]_, >90.5_[boys]_), and obesity (WHO: >97.0; local: >96.8_[girls]_, >95.5_[boys]_; IOTF: >98.6_[girls]_, >98.9_[boys]_). We defined short stature as <3^rd^ percentile of height-for-age.

**RESULTS:** The mean z-scores in LUIS were 0.56 for height, 0.28 for weight, and 0.06 for BMI based on WHO references; 0.15 for height, 0.06 for weight, and -0.01 for BMI based on local references; and 0.19 for BMI based on IOTF references. WHO references provided a worse fit to the LUIS children than local references. Anderson-Darling goodness of fit A^2^ was 578.1 (WHO) vs. 48.1 (local) for height and 124.0 vs. 10.0 for weight, with lower values indicating better fit. WHO (A^2^: 24.3) and local references (A^2^: 0.8) fit the BMI of LUIS children better than IOTF (A^2^: 64.1). The WHO classified fewer children as overweight than the local and IOTF references (WHO: 9% vs. local: 15% vs. IOTF: 13%) but more children as obese (6% vs. 4% vs. 3%). The WHO defined fewer children as being of short stature than the local references (1% vs. 3%).

**CONCLUSIONS:** Our findings suggest that anthropometric data of schoolchildren in Zurich differ notably from WHO and IOTF references potentially leading to misclassification of overweight, obesity, and short stature. Nationally representative and longitudinally collected data are needed to develop new Swiss growth references.

## Introduction

Growth references and their corresponding percentile curves are used by general practitioners and specialist pediatricians to (i) detect abnormal growth patterns that could be related to underlying physical diseases or developmental disorders, (ii) identify short stature or overgrowth, and (iii) determine whether a child fulfills criteria for overweight or obesity based on body mass index (BMI) percentiles. Early identification of atypical growth patterns and developmental deviations is crucial for prompt intervention and management of chronic conditions or developmental delays. For example, short stature can indicate endocrinological, genetic, or chronic systemic diseases. Correct detection of obesity in children is important because childhood obesity is a known predictor of ill health in adulthood and is associated with cardiovascular disease, type 2 diabetes, and cancer [1, 2].

In 2011, the Swiss Society of Pediatrics (pädiatrie schweiz) replaced the growth curves from the Zurich Longitudinal Studies [3] by the WHO curves. For this purpose, two WHO data sets were merged and reanalyzed [4]. Growth standards for children aged 0 to 5 years were based on data from the WHO Multicenter Growth Reference Study conducted between 1997 and 2003 in Brazil, Ghana, India, Norway, Oman, and the USA [5], while growth references for schoolchildren and adolescents aged 5 to 19 years were derived from data collected in the US during the 1960s and 70s [6]. The new growth curves remained a topic of debate. Humans grow taller now than they did several decades ago, and growth varies across ethnicities and regions of the world [7, 8]. Thus, the WHO growth references reflect a different population from the one that lives in Switzerland today and the representativeness of the references has been questioned [9, 10]. In 2019, the Centre for Pediatric Endocrinology Zurich, a privately-run pediatric center in Zurich, created alternative growth references for Swiss children using data collected from pediatric primary care practices in the regions of Zurich and Lucerne [11], referred to here as “local” growth references. The Swiss Society of Pediatrics did not recommend these local growth references because the study was not seen as nationally representative [12]. A third growth reference that is being used in Switzerland was published by the International Obesity Task Force (IOTF) based on data collected between 1963 and 1993 from children in Brazil, Hong Kong, the Netherlands, Singapore, the UK, and the USA [13]. In Switzerland, the IOTF references are mainly used by pediatric endocrinologists to assess overweight and obesity in children.

Given this diversity of growth references used in parallel in Switzerland, and the ongoing debate, we used data from a large population-based study of schoolchildren from the canton of Zurich, the LuftiBus in the school (LUIS) study, to compare the fit of WHO, local, and IOTF growth references to these children. We examined the fit for the entire study population and for subgroups defined by sex, age, origin of parents, socioeconomic status, and degree of urbanization. We also investigated how prevalence estimates of overweight, obesity, and short stature would vary depending on the growth references used to classify the data.

## Materials and methods

### Study design and population

The <u>LuftiBus in the school (LUIS)</u> study is a cross sectional study of lung health in schoolchildren 6–17 years old conducted from 2013 to 2016 in the canton of Zurich (ClinicalTrials.gov NCT03659838, described in detail elsewhere [14]). All 490 schools in the canton of Zurich were invited and 37 schools agreed to participate. Participating schools were visited by a bus—the LuftiBus—equipped with spirometers and scales to measure lung function and anthropometric parameters. The study was approved by the ethics committee of the canton of Zurich (KEK-ZH-Nr: 2014-0491) and informed consent was obtained prior to participation.

### Height, weight, and BMI

For anthropometric measurements, children removed their shoes and wore light clothing. The LuftiBus technicians used a calibrated stadiometer to record body height and a calibrated scale to record body weight. We calculated age and sex dependent z-scores for height, weight, and BMI based on WHO [4], local [11], and IOTF [13] growth references using Cole’s Lambda-Mu-Sigma (LMS) method [15] (see Supplementary Material). We adapted a method from Daymont et al. [16] to detect possible outliers resulting from recording or transcription errors by the LUIS study team: We recentered height, weight, and BMI z-scores around the median z-score at each age year to account for the possibility that the whole study sample might differ from the reference population. We excluded children with a recentered z-score < -4 or > 4 as recommended [17, 18].

We used cutoff values for BMI z-scores published by the WHO, local, and IOTF growth references to classify children into normal weight, overweight, obesity, and severe obesity (Table S6). We defined short stature as <3^rd^ percentile, normal height as between the 3^rd^ and 97^th^ percentiles, and tall stature as >97^th^ percentile for all references [19].

### Potential explanatory factors for growth

We obtained socioeconomic information (origin of parents, education of parents, home address) from parent-completed questionnaires. We grouped parental countries of origin into five categories: Switzerland (both parents born in Switzerland), Switzerland one parent (one parent born in Switzerland, other parent born outside of Switzerland), Northern/Western Europe (both parents born in northern or western Europe), Southern/Eastern Europe (both parents born in southern or eastern Europe), Other/Mixed (parents born in Africa, America, Asia, or Oceania or parents with mixed origin excluding Swiss parents). We defined these geographical regions based on the United Nations standard country or area codes for statistical use, M49 [20]. We classified education of parents into three categories based on the highest education level achieved by one parent: primary education (compulsory schooling only; ≤ 9 years), secondary education (vocational training; 10-13 years), and tertiary education (higher vocational training, college, or university degree; ≥ 14 years). We used the Swiss neighborhood index of socioeconomic position (Swiss-SEP) as an area-based measure of socioeconomic status [21]. The Swiss-SEP ranges from 0 to 100 where a higher value reflects a higher socioeconomic position of that neighborhood. We assigned the nearest Swiss-SEP value to the geographical coordinates of the children’s home address. When the home address was missing (n=405, 11%) we assigned the median Swiss-SEP value of children attending the same school. We classified Swiss-SEP into five categories based on the quintiles published by Panczak et al. [21]. We classified degree of urbanization into three categories based on the location of the school and according to the classification of the Federal Statistical Office [22]: large urban (cities ≥ 50,000 inhabitants and population density ≥ 1500/km^2^), small urban (towns and suburbs ≥ 5000 inhabitants and population density ≥ 300/km^2^), rural (outside of large and small urban regions).

### Statistical analyses

We assessed the fit of the three growth references to the LUIS children by calculating means and standard deviations for height, weight, and BMI z-scores. In a perfectly fitting sample, the z-scores would be standard normally distributed (mean 0, SD 1). We classified mean z-scores deviating >0.5 from the reference population as strong deviation, 0.25-0.5 as moderate deviation, 0.05-0.25 as weak deviation, and 0-0.05 as no noticeable deviation [8]. We also used the Anderson-Darling test to evaluate the fit of the z-score distributions to the expected standard normal distribution. It quantifies the goodness of fit, measured as A^2^, where a lower value reflects a better fit. The p-value is the probability for the null hypothesis that the z-score distribution is drawn from a standard normal distribution.

We assessed the fit of growth references in the entire population and within strata defined by factors selected a priori based on a literature review and expert opinion: sex, age, origin of parents, Swiss-SEP, and degree of urbanization. For this we estimated mean z-scores by age and their pointwise 95% confidence interval (CI) using penalized cubic (3^rd^ degree) B-spline regression over age for girls and boys separately to assess the fit of the growth references graphically across age [23]. We compared the fit of the growth references across potential explanatory factors again by computing the Anderson-Darling A^2^. Furthermore, we used univariable linear regression models to determine the association between potential explanatory factors and z-scores of height, weight, and BMI derived from the local references. We included variables jointly in multivariable models based on significance in univariable models (p <0.05) and measures of model fit.

We calculated prevalence of BMI and height categories and their 95% CI based on the different references. We used Cohen’s Kappa to quantify agreement between growth references. We interpreted Cohen’s Kappa 0.41-0.60 as moderate, 0.61-0.80 as substantial, and 0.81-1.00 as almost perfect agreement according to Landis and Koch [24]. We repeated prevalence estimates of BMI and height categories in strata defined by sex, age, origin of parents, Swiss-SEP, and degree of urbanization.

We used the statistical software R Studio version 2023.3.1 [25] with the R version 4.2.0 [26] for all analyses.

## Results

### Study population

Of 3870 eligible children from the LUIS study, we excluded 107 children with missing height or weight measurements, and a further eight children with height, weight, or BMI z-scores of < -4 or > 4 (Figure S1), leaving 3755 children (50% girls) in our analysis. For analyses requiring origin of parents, we further excluded 390 (10%) children with unknown origin of parents. The median age was 13 years (IQR 10-14, Table 1 and Table S1). Most children had parents originally from Switzerland (both parents: 51%, one parent: 16%). The median Swiss-SEP was 69 (IQR 61-76), and half of the children were in the highest Swiss-SEP quintile category. Boys and girls had similar median height and weight.

**Table 1:** Characteristics of participants of the <u>LuftiBus in the School (LUIS)</u> study.

|  | **Girls, n (%)**<br>1872 (50) | **Boys, n (%)**<br>1883 (50) | **Total, n (%)**<br>3755 (100) |
| --- | --- | --- | --- |
| **Age (years)** |  |  |  |
| median (IQR) | 12.8 (10.1, 14.2) | 12.7 (9.9, 14.3) | 12.8 (10.0, 14.3) |
| groups , n (%) |  |  |  |
| 6-8 | 333 (18) | 345 (18) | 678 (18) |
| 9-11 | 441 (24) | 440 (23) | 881 (23) |
| 12-14 | 838 (45) | 825 (44) | 1663 (44) |
| 15-17 | 260 (14) | 273 (14) | 533 (14) |
| **Origin of parents, n (%)** |  |  |  |
| Switzerland (both parents) | 965 (52) | 968 (51) | 1933 (51) |
| Switzerland (one parent) | 279 (15) | 305 (16) | 584 (16) |
| Northern/Western Europe (both parents) | 103 (6) | 100 (5) | 203 (5) |
| Southern/Eastern Europe (both parents) | 199 (11) | 183 (10) | 382 (10) |
| Other/Mixed | 142 (8) | 121 (6) | 263 (7) |
| Missing | 184 (10) | 206 (11) | 390 (10) |
| **Education of parents, n (%)** |  |  |  |
| Primary education | 54 (3) | 51 (3) | 105 (3) |
| Secondary education | 475 (25) | 473 (25) | 948 (25) |
| Tertiary education | 714 (38) | 679 (36) | 1393 (37) |
| Missing | 629 (34) | 680 (36) | 1309 (35) |
| **Swiss-SEP 2.0\*** |  |  |  |
| median (IQR) | 69 (61, 76) | 69 (62, 77) | 69 (61, 76) |
| groups, n (%) |  |  |  |
| 1: 0 - 49 | 70 (4) | 68 (4) | 138 (4) |
| 2: 50 - 56 | 232 (12) | 227 (12) | 459 (12) |
| 3: 57 - 61 | 221 (12) | 210 (11) | 431 (11) |
| 4: 62 - 69 | 405 (22) | 436 (23) | 841 (22) |
| 5: 70 - 100 | 944 (50) | 942 (50) | 1886 (50) |
| **Urbanization, n (%)** |  |  |  |
| Rural | 128 (7) | 119 (6) | 247 (7) |
| Small urban: towns, suburbs | 807 (43) | 828 (44) | 1635 (44) |
| Large urban: cities | 937 (50) | 936 (50) | 1873 (50) |
| **Height (cm)** |  |  |  |
| median (IQR) | 156 (141, 164) | 156 (141, 170) | 156 (141, 166) |
| **Weight (kg)** |  |  |  |
| median (IQR) | 45 (33, 56) | 44 (33, 58) | 45 (33, 57) |

\* Swiss-SEP: Swiss socioeconomic position version 2.0, census data from 2012 - 2015. The 5 groups are based on quintiles published by Panczak et al. [21]. A higher score relates to a higher socioeconomic position.
IQR: Inter quartile range

### Fit of WHO, local, and IOTF references to children in LUIS

LUIS children deviated strongly in height (mean z-score: 0.56), moderately in weight (0.28), but only weakly in BMI (0.06) from WHO references according to the criteria of Natale et al. [8] (Table 2). Mean height (0.15) and weight (0.06) deviated weakly, while BMI (-0.01) did not deviate noticeably from local references. Mean BMI (0.19) deviated weakly from IOTF references.

**Table 2:**
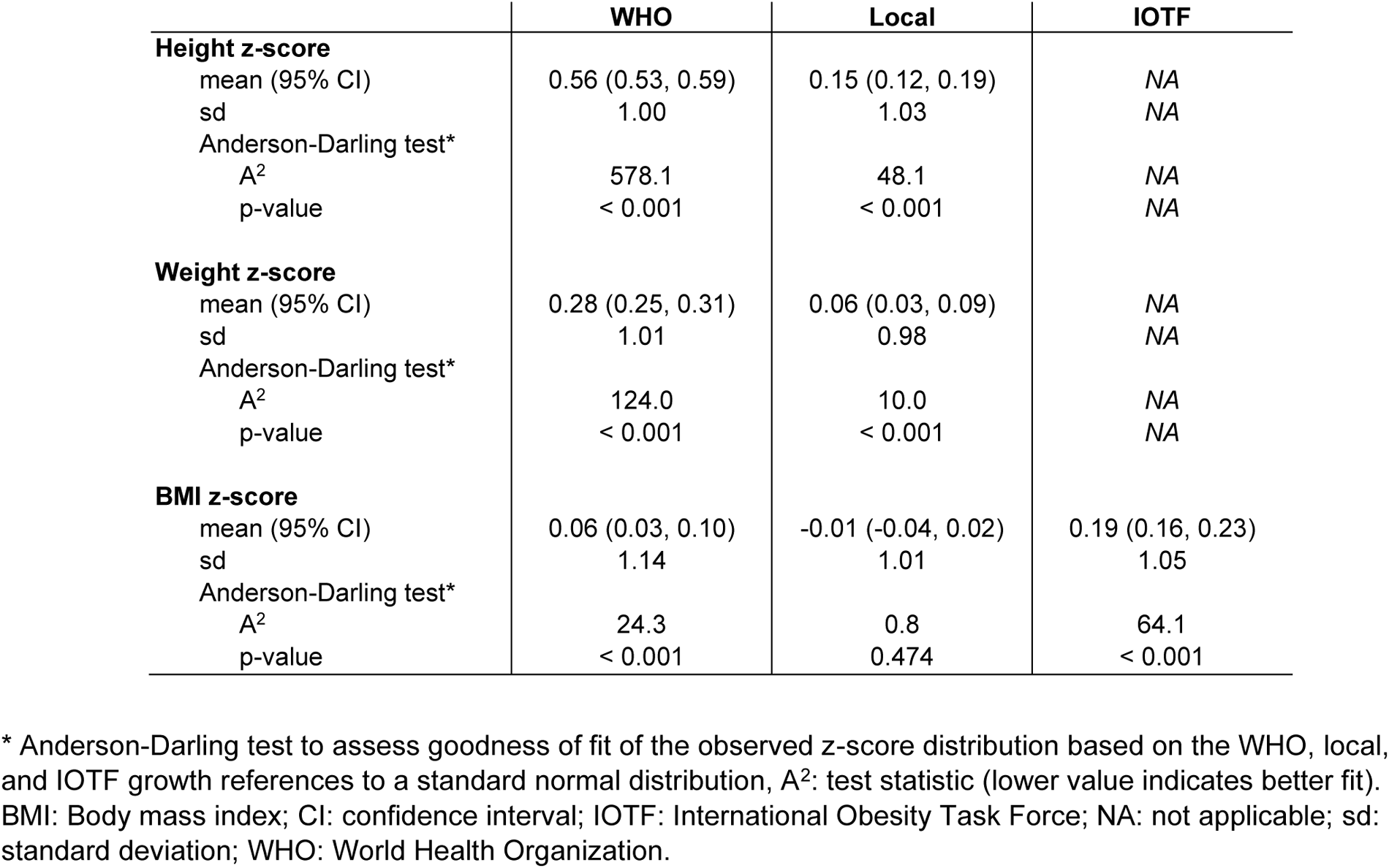
Mean height, weight, and BMI z-scores of 3755 participants of the <u>LuftiBus in the School (LUIS)</u> study based on the WHO, Local, and IOTF references; and the fit of the z-scores to a standard normal distribution.

|  | WHO | Local | IOTF |
| --- | --- | --- | --- |
| **Height z-score** |  |  |  |
| mean (95% CI) | 0.56 (0.53, 0.59) | 0.15 (0.12, 0.19) | NA |
| sd | 1.00 | 1.03 | NA |
| Anderson-Darling test* |  |  |  |
| A<sup>2</sup> | 578.1 | 48.1 | NA |
| p-value | < 0.001 | < 0.001 | NA |
| **Weight z-score** |  |  |  |
| mean (95% CI) | 0.28 (0.25, 0.31) | 0.06 (0.03, 0.09) | NA |
| sd | 1.01 | 0.98 | NA |
| Anderson-Darling test* |  |  |  |
| A<sup>2</sup> | 124.0 | 10.0 | NA |
| p-value | < 0.001 | < 0.001 | NA |
| **BMI z-score** |  |  |  |
| mean (95% CI) | 0.06 (0.03, 0.10) | -0.01 (-0.04, 0.02) | 0.19 (0.16, 0.23) |
| sd | 1.14 | 1.01 | 1.05 |
| Anderson-Darling test* |  |  |  |
| A<sup>2</sup> | 24.3 | 0.8 | 64.1 |
| p-value | < 0.001 | 0.474 | < 0.001 |

\* Anderson-Darling test to assess goodness of fit of the observed z-score distribution based on the WHO, local, and IOTF growth references to a standard normal distribution, A<sup>2</sup>: test statistic (lower value indicates better fit). BMI: Body mass index; CI: confidence interval; IOTF: International Obesity Task Force; NA: not applicable; sd: standard deviation; WHO: World Health Organization.

According to the Anderson-Darling test, the local height references (A^2^: 48.1) fit better than the WHO references (A^2^: 578.1) (Table 2 and Figure 1). The distribution of height measurements among LUIS children deviated significantly from both reference populations (test for standard normal distribution of z-scores, p <0.001). The fit to body weight was better for local (A^2^: 10.0) than for WHO references (A^2^: 124.0), but neither of the z-scores followed the standard normal distribution (p <0.001). The fit to BMI was better for local (A^2^: 0.8) than for WHO (A^2^: 24.3) and IOTF references (A^2^: 64.1). Only the local BMI z-scores agreed with the standard normal distribution (p = 0.474).

**Figure 1.**
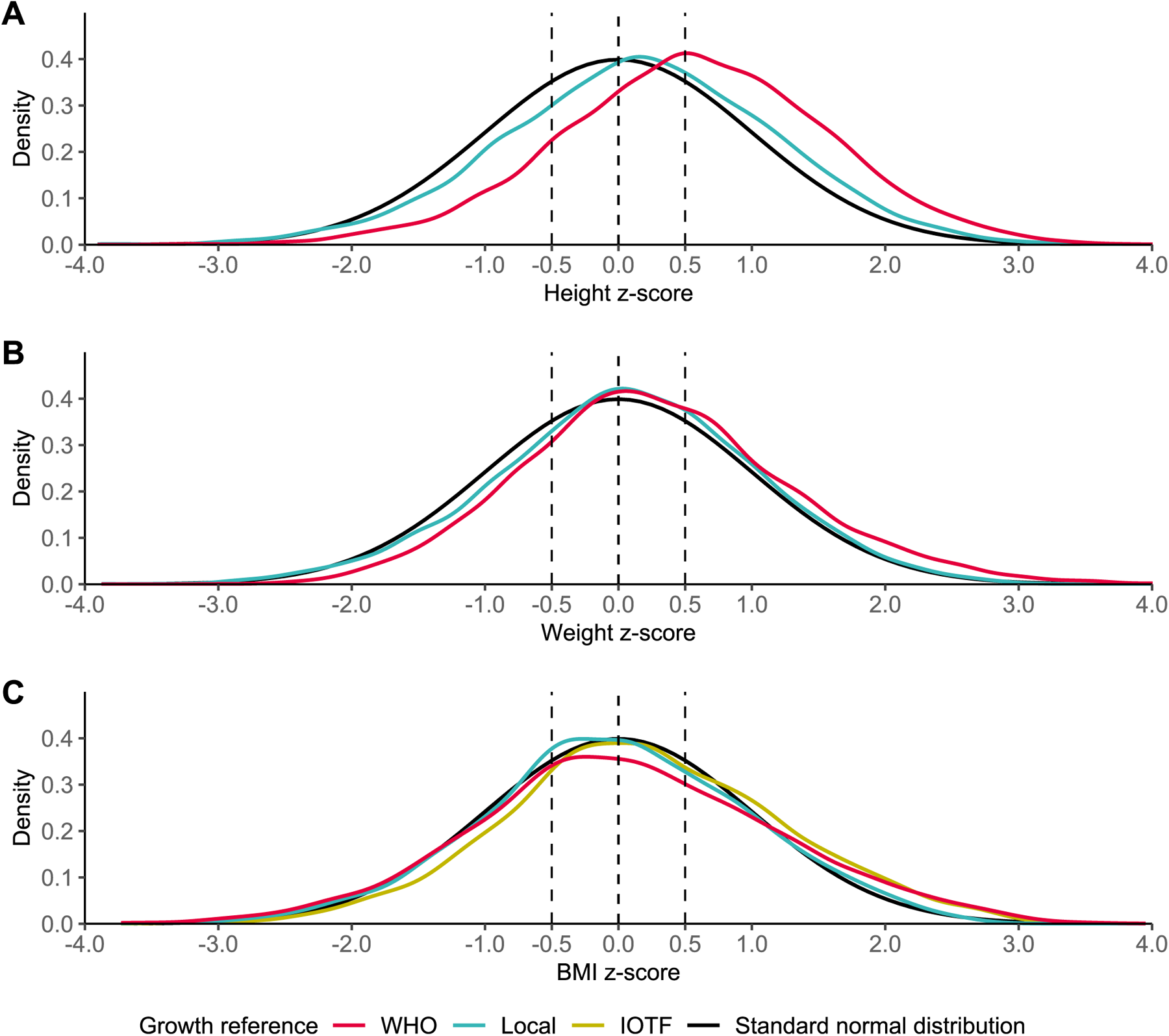
Z-score distributions of **A)** height, **B)** weight, and **C)** BMI for 3755 participants of the <u>LuftiBus in the</u> <u>school (LUIS)</u> study. Z-score distributions based on WHO, local, and IOTF references are shown versus a standard normal distribution (perfect fit). BMI: Body Mass Index; IOTF: International Obesity Task Force; WHO: World Health Organization.

A better fit of local compared to WHO references was also apparent from age-specific mean z-scores estimated by penalized cubic B-spline regression (Figure 2 and Figure S2). Mean height and weight z-scores based on local references were closer to the expected zero value compared to WHO references for most ages in both boys and girls. Only among girls 17 years old, for height, and 10–11 years old, for weight, were the WHO mean z-scores closer to zero than z-scores derived from local references, although 95% CIs of mean z-scores from WHO and local references overlapped. Height z-scores were closer to zero among older children (>10 years) than among younger children (<8 years) for both WHO and local references. Mean BMI z-scores deviated only weakly from zero in both boys and girls for all three references. IOTF BMI z-scores were further away from zero than the WHO and local ones in older children (>12 years).

**Figure 2.**
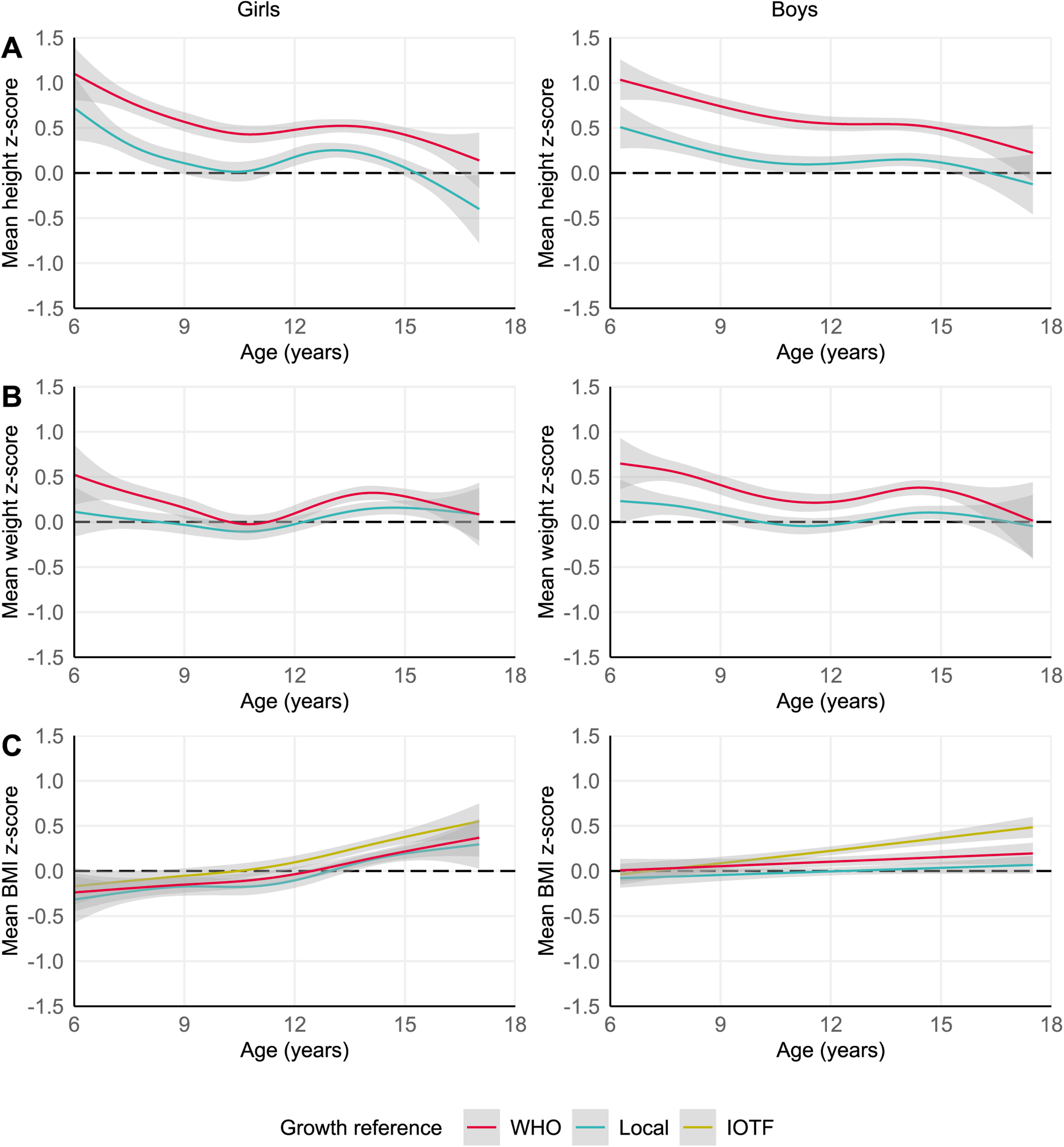
Mean **A)** height, **B)** weight, and **C)** BMI z-scores over age for 3755 participants of the <u>LuftiBus in the</u> <u>school (LUIS)</u> study split by sex. Mean z-scores based on WHO, local, and IOTF references and their 95% confidence intervals were locally estimated by penalized cubic B-spline regression on age. BMI: Body Mass Index; IOTF: International Obesity Task Force; WHO: World Health Organization.

### Factors influencing fit of growth references

Local references fit height and weight of LUIS children better than WHO references across all strata of sex, age, origin of parents, and Swiss-SEP, and in most categories of urbanization (Figure S3 and Table S2). Local references fit BMI best in girls and boys and in most categories of age, origins of parents, Swiss-SEP, and degree of urbanization.

Univariable and multivariable linear regression confirmed that z-scores of height, weight, and BMI based on local references differ by age, origin of parents, Swiss-SEP, and degree of urbanization (Tables S3, S4, and S5). Height z-scores tended to decrease with age and increase with Swiss-SEP. Height z-scores were larger in children with parents born in Northern/Western Europe and Southern/Eastern Europe compared to children with parents born in Switzerland. Height z-scores were larger in children living in urban regions compared to rural regions. Weight and BMI z-scores tended to decrease with Swiss-SEP. Weight and BMI z-scores were higher in children with parents from Southern/Eastern Europe and mixed origin compared to children with parents from Switzerland. Weight and BMI z-scores were higher in children living in an urban compared to a rural region. The linear regression did not show any differences in height, weight, or BMI z-scores by sex.

### Prevalence of overweight, obesity, and short stature differ by growth reference

Cohen’s Kappa showed substantial—but not perfect—agreement of the classification into BMI categories between the different growth references (Table S6). The prevalence of normal weight in LUIS children was comparable between WHO and IOTF (85%), but slightly lower for local references (81%, Figure 3). The WHO classified fewer children as overweight (8%) compared to local (15%) and IOTF (13%) references. In contrast, the WHO classified more children as obese (5%) and severely obese (2%) compared to IOTF (obese 3%, severely obese: 0.3%), and more children as severely obese compared to local references (0.6%).

**Figure 3.**
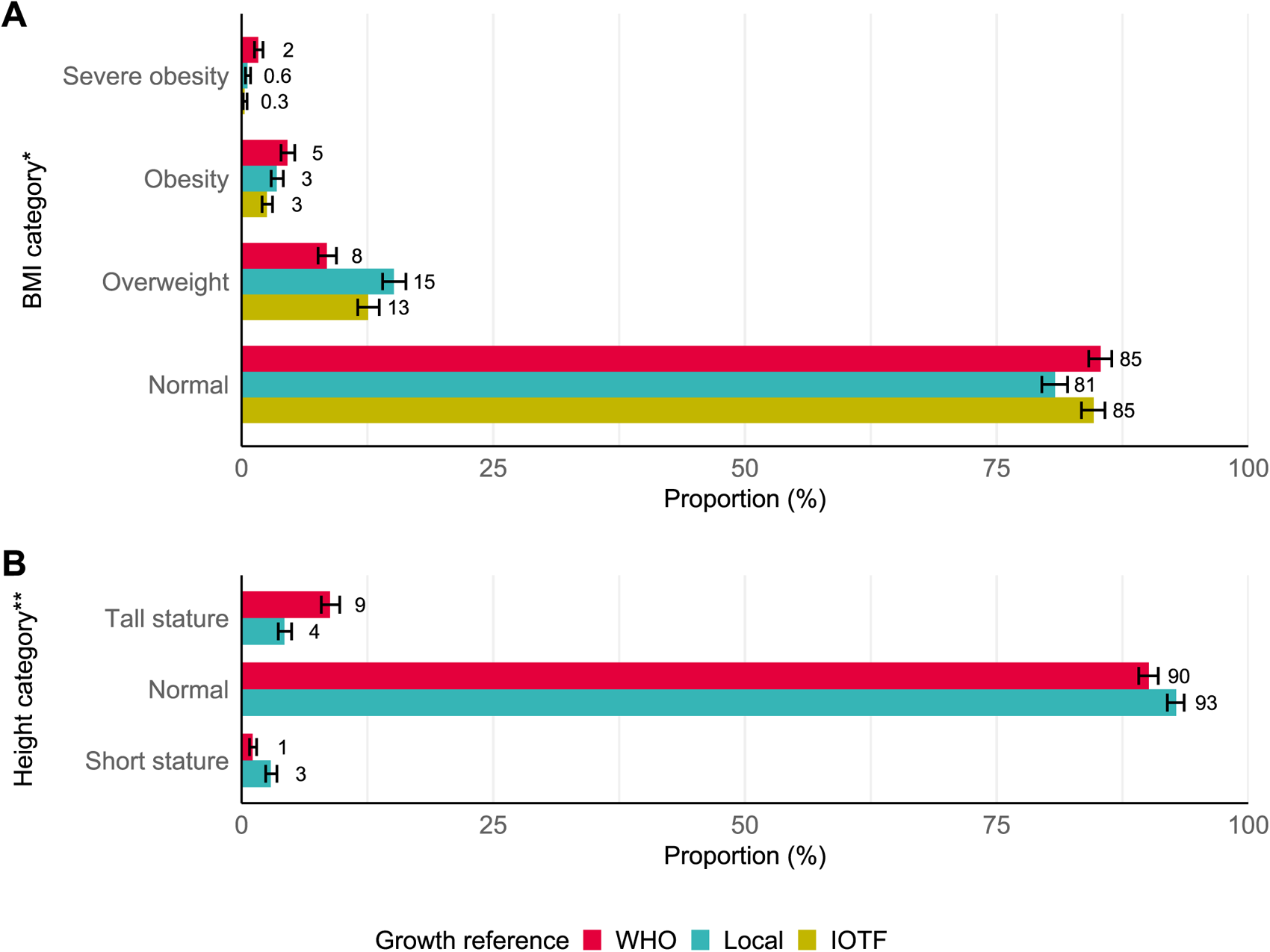
Prevalence of (A) BMI and (B) height categories and their 95% CIs in 3755 participants of the <u>LuftiBus in the school (LUIS)</u> study. * BMI categories were defined based on cutoff values of the WHO, local, and IOTF, supplementary Table S6. ** Height categories were defined by percentiles: short stature (<3^rd^), normal (3^rd^≤≥97^th^), and tall stature (>97^th^). BMI: Body Mass Index; CI: confidence interval IOTF: International Obesity Task Force; WHO: World Health Organization.

BMI categories differed across origin of parents, Swiss-SEP, and degree of urbanization, but varied little between age groups or between boys and girls (Figure S4). Overweight, obesity, and severe obesity were more prevalent in children whose parents were of Southern/Eastern Europe and Other/Mixed origin compared to those of Swiss origin. We observed a higher prevalence of overweight, obesity, and severe obesity in the two lowest Swiss-SEP categories compared to the highest Swiss-SEP category. Overweight was more prevalent in large and small urban compared to rural areas. The classification into BMI categories by the three growth references had a substantial agreement in all categories (Table S7).

The height categories differed between WHO and local references, which was reflected by only moderate agreement according to Cohen’s Kappa (Figure 3 and Table S6). The WHO estimated fewer children as being of short stature (1%) compared to local references (3%) but estimated more children as being of tall stature (9% vs. 4%). The height categories did not differ across sex, age group, origin of parents, Swiss-SEP, or degree of urbanization (Figure S4).

## Discussion

Comparing anthropometric data from LUIS, a population-based study of schoolchildren from the canton of Zurich, to three currently used growth references in Switzerland, this study found a poor fit of WHO and IOTF growth references, while local references, developed recently in the same region, Zurich and Luzern, fit better overall and within population strata. Children in LUIS were taller compared to the WHO reference population, particularly in younger ages. Children living in urban environments and those whose parents came from Northern or Western Europe were taller compared to children from rural regions and with Swiss parents. Children from urban regions, lower Swiss-SEP index, and whose parents came from Southern or Eastern Europe had a higher BMI. Prevalence estimates for being overweight, obese, or of short stature differed depending on the growth references that were used to classify children.

### Comparison with other studies

The anthropometric data of children from the Zurich region differed from the WHO growth references as previously reported for this region [11]. The same has been reported in Germany [27], Austria [28], and France [29]. Improved socioeconomic and nutritional circumstances over the last century have contributed to a general increase in adult human stature across the world [7, 30], and this has been accompanied by a decrease in the age of onset of puberty [31]. Since the WHO growth references for schoolchildren and adolescents used data collected in the 1960s and 1970s [32], both an increased adult target height and an earlier onset of puberty could explain why the children in our study (2013-2016) were taller, particularly at younger ages. Height is strongly determined by genetic background with twin studies suggesting up to 80% heritability of body stature [33]. Adult men and women of Central and Latin American or Asian origin tend to be shorter compared to North Europeans, Central Europeans, or North Americans, with a gap of up to 20 cm between the tallest and shortest populations [7]. The US study from which WHO references were derived consisted of children with white (85%) and African-American (15%) ethnicity [34]. In our study, 80% of the children were of European origin and 20% of mixed or unknown origin. Such differences in ethnic mix can also influence the fit of growth references. Our multivariable regression models confirmed parental origin, area-based socioeconomic status, and urbanization as significant predictors of height, weight, and BMI although the models explained little of the overall variability between study participants since the adjusted R^2^ for the regression model was 1.3% for height, 4.4% for weight, and 5.5% for BMI.

Children in LUIS were slightly taller than the population from which the local growth references were derived. LUIS included more children with a Swiss origin and fewer children with another/mixed background than the local reference population (Table S1), but this seems not to be the reason because height z-scores were comparable between Swiss children and those of Other/Mixed origin. However, the slightly higher socioeconomic status of children in LUIS compared to the local reference population might have contributed to the small height difference [35].

We found a clear difference in prevalence estimates of obesity depending on whether we used WHO, local, or IOTF references to classify children. This has also been reported for IOTF and US Centers for Disease Control and Prevention (CDC) references [36]. Prevalence of obesity (3%) and overweight (13%) in LUIS, using IOTF references, were almost identical to reports of Gesundheitsförderung Schweiz: 4% obesity and 13% overweight in schoolchildren (6-16 years) in the city of Zurich for the years 2014/15 [37], which also were calculated with IOTF references. Similar to other Swiss studies [38, 39], we found that children living in lower Swiss-SEP locations, in urban areas, and those whose parents originated from Southern or Eastern Europe had a higher prevalence of overweight and obesity.

### Strengths and limitations

Our study is limited by regionality, by selection bias of high socioeconomic status in the study population, and by lack of information on pubertal stages. We cannot extrapolate our results to all Swiss children because LUIS only includes children from the canton of Zurich, in German-speaking Switzerland. More people have a migration background in Zurich (46%) than in Switzerland as a whole (40%) [40]. Half of the study participants lived in a location in the highest Swiss-SEP quintile. Although the Swiss-SEP in LUIS (median 69, IQR 61-76) was only slightly higher than for all households in Zurich with one or more children (median 66, IQR 58-73) [14], this might differ from other regions and could have led to an overestimation of height and an underestimation of BMI in our study resulting in poorer fit for height references and in better fit for BMI references. We did not have information on pubertal Tanner stage and could not account for puberty in the analysis. An earlier onset of the pubertal growth spurt compared to the reference populations could explain why the excess height was particularly evident in the youngest age group of 6-9-year-olds.

The study benefits from the population-based approach, the standardized anthropometric measurements, and the incorporation of information on origin of parents and socioeconomic status.

### Implications

This study confirms that WHO references currently used in Switzerland fit the local population poorly—or at least schoolchildren in the Zurich region. Poorly fitting growth references can lead to misclassification of overweight, obesity, or short stature, resulting in uncertainties in clinical decision making that can include delayed treatments and unnecessary referrals [41, 42]. Pediatricians should mainly rely on longitudinal trajectories of height and weight to detect abnormal growth rather than on single measurements and cutoff values [43]. Hence, new growth references representing children living in Switzerland are needed.

New Swiss growth references should be derived from nationally representative data. Such data should include children living in German-, French-, and Italian-speaking regions of Switzerland so that new growth references can be used by pediatricians and general practitioners [4, 10, 12]. Longitudinally collected data from primary care, hospitals (SwissPedHealth, https://www.swisspedhealth.ch), and school medical services could contribute to new evidence-based growth references.

In conclusion, the growth references currently used in Switzerland may lead to misclassification of overweight, obesity, and short stature. Children would benefit from new Swiss growth references, which will require nationally representative and longitudinally collected data.

### Availability of data and material

Researchers can obtain datasets for analysis if a detailed concept sheet is presented for the planned analyses and approved by the principal investigators (Alexander Moeller, Philipp Latzin and Claudia Kuehni).

## Acknowledgments

We thank the school teams and the families for participating in the study. We thank the field workers and study personnel for conducting the study. We thank Claudia Berlin and the Swiss National Cohort (SNC, www.swissnationalcohort.ch) for providing data on the Swiss-SEP for the canton of Zurich. We thank the Swiss Federal Statistical Office for providing data on schools and degree of urbanization of municipalities in the canton of Zurich.

The LuftiBus in the School (LUIS) study group: Alexander Moeller, Jakob Usemann, Florian Singer (Department of Respiratory Medicine, University Children’s Hospital Zurich and Childhood Research Centre, University of Zurich, Switzerland); Philipp Latzin, and Johanna M. Kurz (Division of Paediatric Respiratory Medicine and Allergology, Dept of Paediatrics, Inselspital, Bern University Hospital, University of Bern, Switzerland); Claudia E. Kuehni, Rebeca Mozun, Cristina Ardura-Garcia, Myrofora Goutaki, Eva S.L. Pedersen and Maria Christina Mallet (Institute of Social and Preventive Medicine, University of Bern, Switzerland); and Kees de Hoogh (Swiss Tropical and Public Health Institute, Basel, Switzerland).

