## Supplementary material for "Evaluating Swiss growth reference curves: A comparative analysis in Zurich schoolchildren"

### 1 Calculation of z-scores

We calculated age and sex dependent z-scores for body height, body weight, and BMI based on WHO [1], local [2], and IOTF [3] growth references. We used cubic interpolation of the published LMS-values to determine the exact LMS-values for each age in days, as described by Vidmar, Cole and Pan [4]. We used a formula from Cole and Green [5] to calculate z-scores (z) with our measurement ($x$) and the L($\lambda$), M($\mu$), and S($\sigma$) values, see formula (1):

$$z = \frac{\left(\frac{x}{\mu}\right)^{\lambda} - 1}{\lambda\sigma} \tag{1}$$

We used a correction method described by the WHO to correctly compute z-scores >3 and <-3 [6], see formulas (2), (3), (4) ,(5) and (6):

$$z^{*} = \begin{cases} z & if \quad |z| \leq 3 \\ 3 + \left(\frac{x - SD3pos}{SD23pos}\right) & if \quad z > 3 \\ -3 + \left(\frac{x - SD3neg}{SD23neg}\right) & if \quad z < -3 \end{cases} \tag{2}$$

where

$$SD3pos = \mu(1 + \lambda * \sigma * 3)^{\frac{1}{\lambda}} \tag{3}$$

$$SD3neg = \mu\big(1 + \lambda * \sigma * (-3)\big)^{\frac{1}{\lambda}} \tag{4}$$

$$SD23pos = \mu(1 + \lambda * \sigma * 3)^{\frac{1}{\lambda}} - \mu(1 + \lambda * \sigma * 2)^{\frac{1}{\lambda}} \tag{5}$$

$$SD23neg = \mu\big(1 + \lambda * \sigma * (-2)\big)^{\frac{1}{\lambda}} - \mu\big(1 + \lambda * \sigma * (-3)\big)^{\frac{1}{\lambda}} \tag{6}$$

We adapted a method from Daymont et al. [7] to detect possible outliers resulting from recording or transcription errors by the LUIS study team: We recentered height, weight, and BMI z-scores around the median z-score at each age year to account for the possibility that the whole study sample could be different from the reference population. We excluded children with a recentered z-score of <-4 or >4 [8, 9]. We used the recentered z-scores only for data cleaning.

### 2 Supplementary Figures and Tables

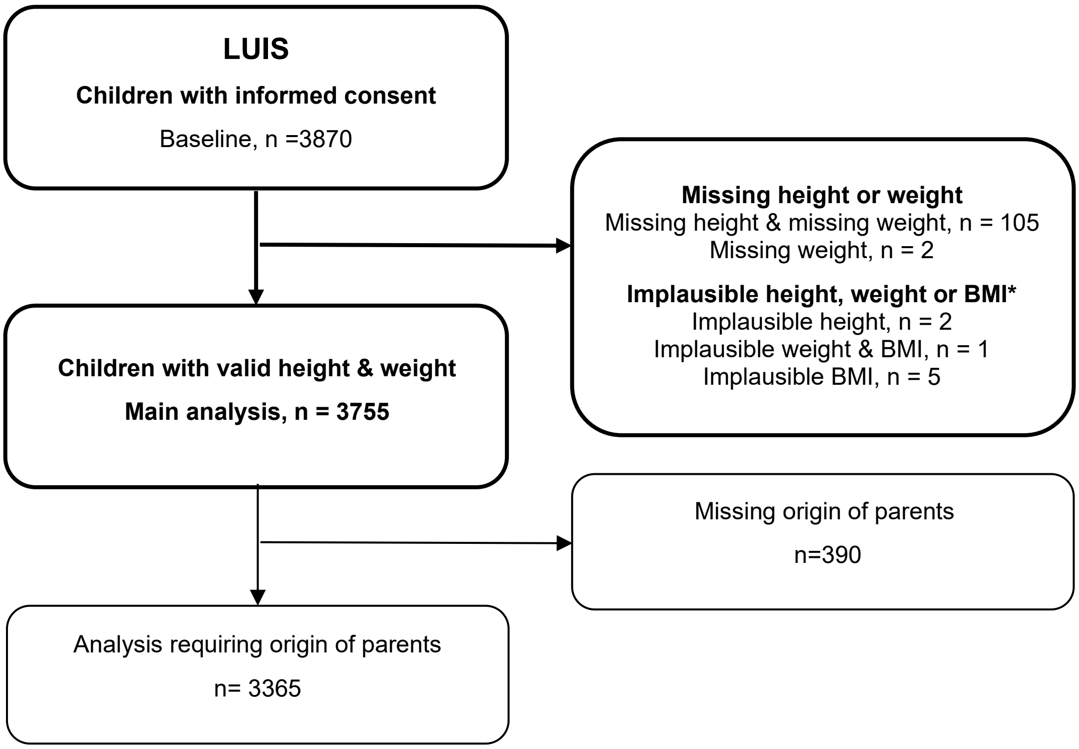


**Figure S1.** Flow diagram of the study population.
* We excluded 8 children with recentered height, weight, or BMI z-scores <-4 or >4.

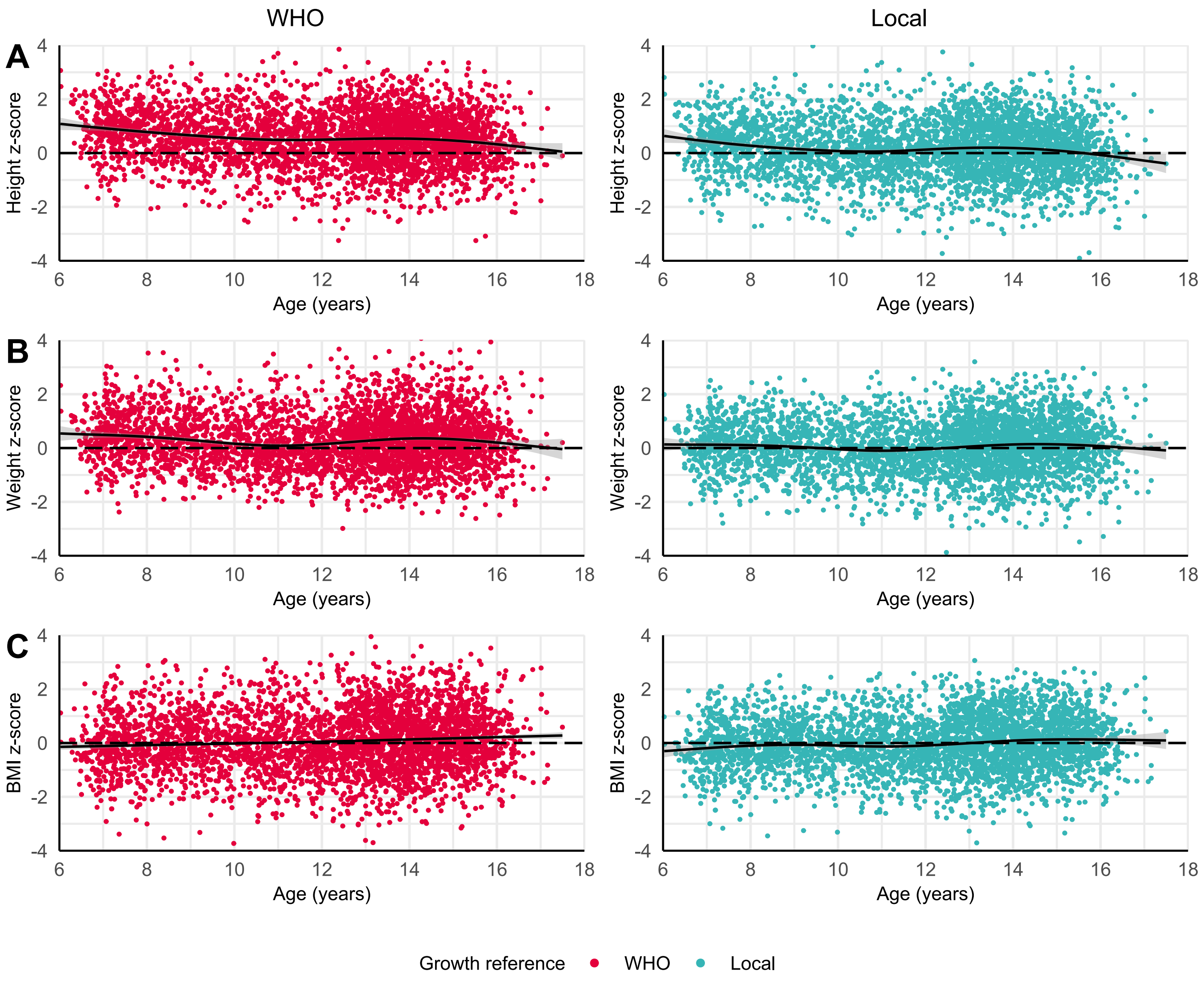


**Figure S2.** Scatterplot of **A)** height, **B)** weight, and **C)** BMI z-scores by age for 3755 children of the LuftiBus in the School (LUIS) study based on WHO and local references. Mean z-scores and their 95% confidence intervals were locally estimated with penalized cubic B-Spline regression.
BMI: Body mass index; WHO: World Health Organization.

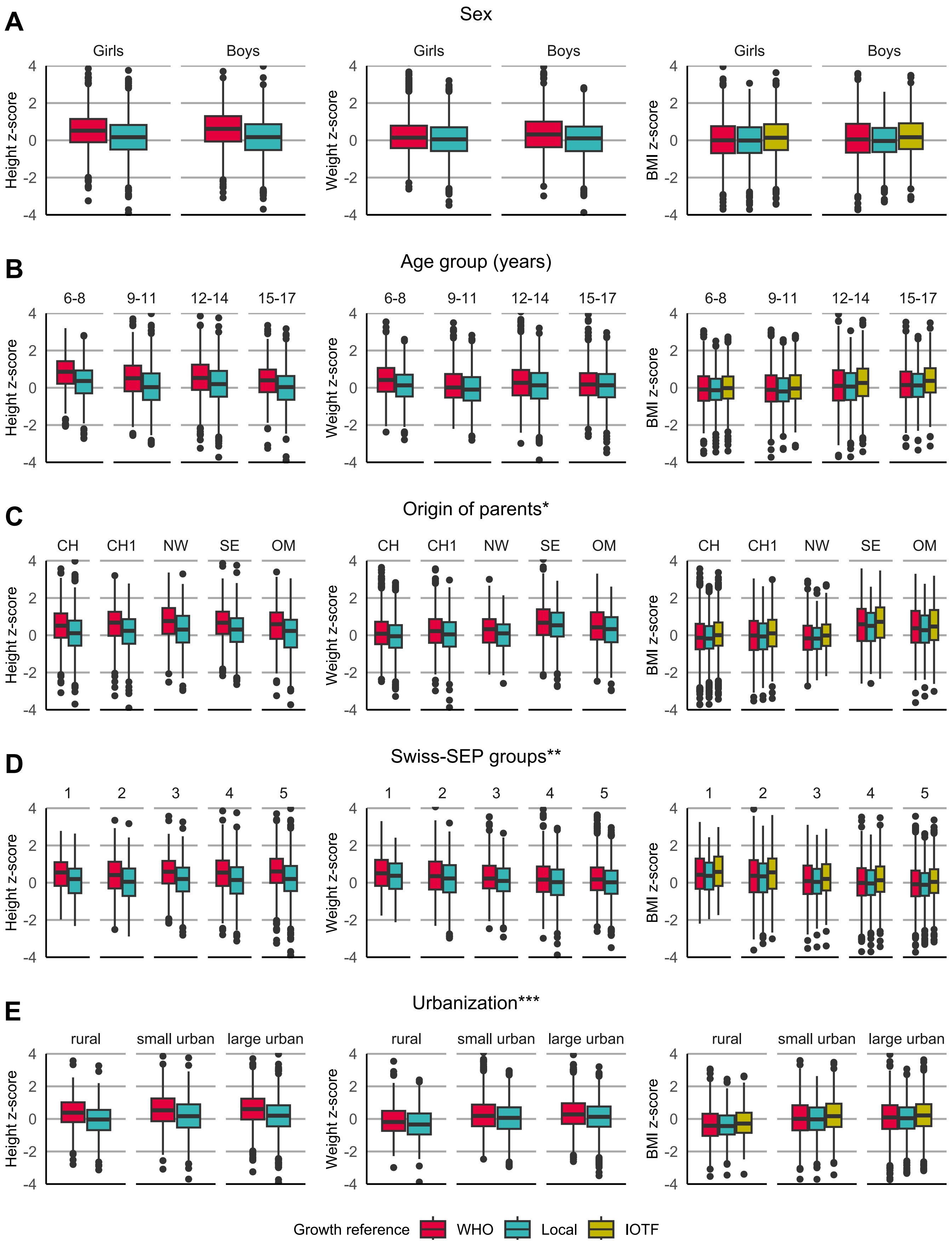


**Figure S3.** Height, weight, and BMI z-scores of 3755 children of the LuftiBus in the School (LUIS) study based on WHO, local, and IOTF references by **A)** sex, **B)** age, **C)** origin of parents, **D)** Swiss-SEP, and **E)** urbanization.
* Origin of parents: CH: both parents from Switzerland; CH1: one parent from Switzerland; NW: both parents from Northern/Western Europe; SE: both parents from Southern/Eastern Europe; OM: Other/Mixed, parents from other or not from the same regions. For the origin of parents, we included n = 3365 children.
** Swiss-SEP: Swiss socioeconomic position version 2.0, census data from 2012 - 2015. The 5 groups are based on quintiles published by Panczak et al. [10]. A higher score relates to a higher socioeconomic position.
*** Urbanization: rural: rural areas; small urban: towns and suburbs; large urban: cities.
BMI: Body Mass Index; WHO: World Health Organization. IOTF: International Obesity Task Force

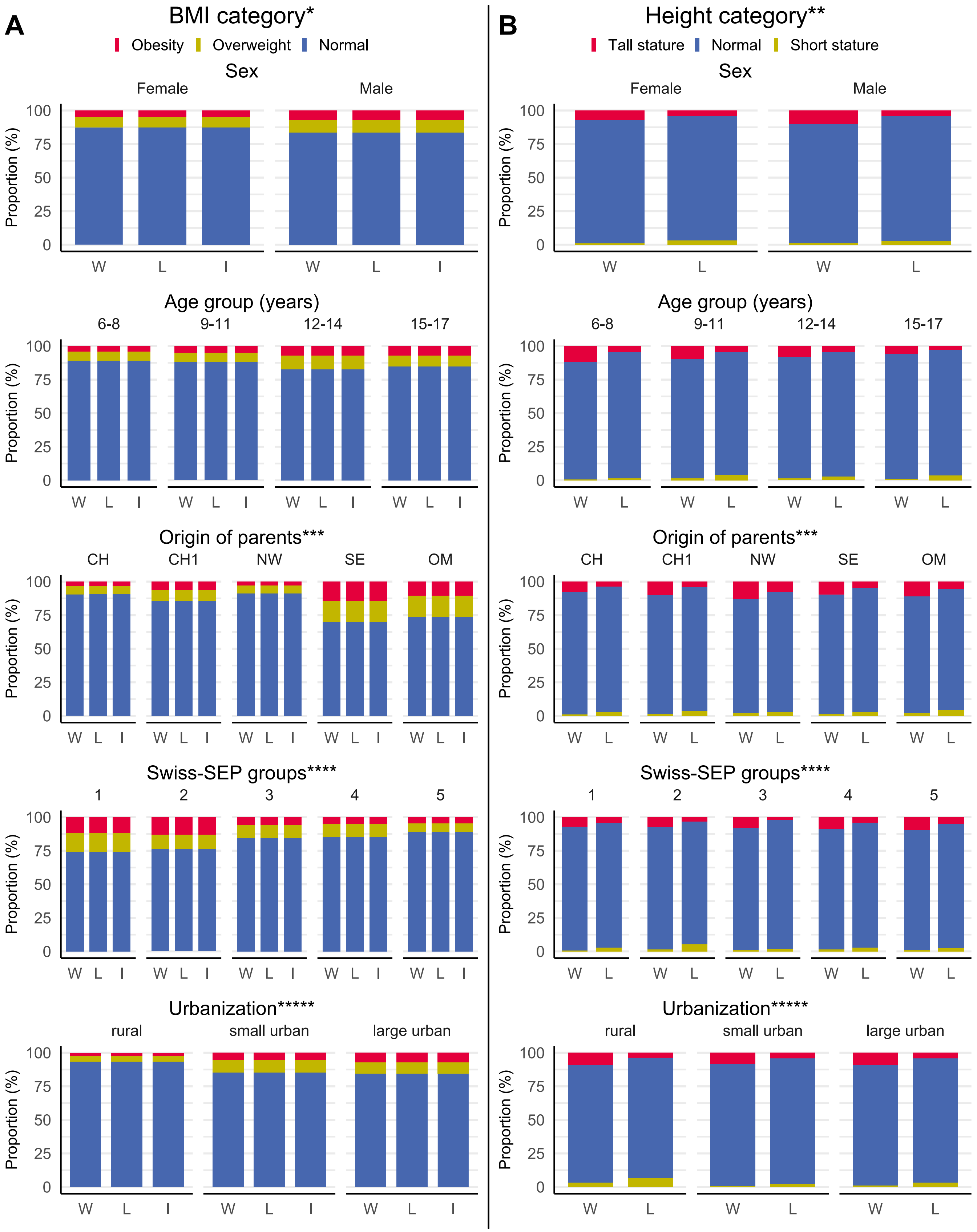


**Figure S4.** Prevalence of the **A)** BMI and **B)** height categories for 3755 children of the LuftiBus in the school (LUIS) study by sex, age, origin of parents, Swiss-SEP, and urbanization.
* BMI categories were defined based on cutoff values of the WHO, of local, and IOTF, see supplementary Table S1. For readability we plotted obesity (obesity + severe obesity), overweight, and normal weight.
** Height categories were defined by percentiles: short stature (<3rd), normal (3rd≤≥97th), and tall stature (>97th).
*** Origin of parents: CH: both parents from Switzerland; CH1: one parent from Switzerland; NW: both parents from Northern/Western Europe; SE: both parents from Southern/Eastern Europe; OM: Other/Mixed, parents from other or not from the same regions. For the origin of parents, we included n = 3365 children.
**** Swiss-SEP: Swiss socioeconomic position version 2.0, census data from 2012 - 2015. The 5 groups are based on quintiles published by Panczak et al. [10]. A higher score relates to a higher socioeconomic position.
***** Urbanization: rural: rural areas; small urban: towns and suburbs; large urban: cities.
BMI: Body Mass Index; L: local; W: WHO, World Health Organization; I: IOTF, International Obesity Task Force.

**Table S1:**
Characteristics of children of the LuftiBus in the School (LUIS) study compared to the population of the local growth references.

| | **LUIS Children N = 3755** | **Local N = 30,141** |
|---|---|---|
| **Sex**, n (%) | | |
| Girls | 1872 (50) | 16,540 (55) |
| Boys | 1883 (50) | 13,601 (45) |
| **Age (years)** | | |
| groups, n (%) | | |
| 6-7 | 409 (11) | 2166 (18)** |
| 8-9 | 533 (14) | 1327 (11) |
| 10-11 | 617 (16) | 3528 (29) |
| 12-13 | 1,076 (29) | 1710 (14) |
| 14-15 | 1,019 (27) | 1983 (16) |
| 16-17 | 101 (3) | 1514 (12) |
| **Origin of parents***, n (%) | | |
| CH | 1,933 (51) | 9510 (32) |
| CH (one parent) | 584 (16) | 3674 (12) |
| DE, FR, AUT, SCAND, BENELUX | 184 (5) | 1265 (4) |
| IT, ES, PT | 81 (2) | 1176 (4) |
| BALKAN | 257 (7) | 2008 (7) |
| Turkey | 18 (<1) | 433 (1) |
| Other/Mixed/Missing | 698 (19) | 12,075 (40) |
| **Swiss-SEP 2.0***** | | |
| Observed quartiles | | |
| 1 | <61.4 | <58.8 |
| 2 | 61.4 – 69.5 | 58.8 – 64.9 |
| 3 | 69.5 – 76.5 | 64.9 – 72.3 |
| 4 | >76.5 | >72.3 |

* Origin of parents: CH (both parents): both parents from Switzerland; CH (one parent): one parent from Switzerland; DE, FR, AUT, SCAND, BENELUX: both parents from Germany, France, Austria, Denmark, Norway, Finland, Sweden, Island, Belgium, the Netherlands, or Luxembourg; IT, ES, PT: both parents from Italy, Spain, or Portugal; BALKAN: both parents from Albania, Bosnia and Herzegovina, Bulgaria, Croatia, Kosovo, Montenegro, North Macedonia, Romania, Serbia, or Slovenia; Turkey: both parents from Turkey; Other/Mixed/Missing: parents from other or not from the same regions, or missing origin of parents.
** The population of the local growth references also included 14,784 children below 6 years and 3129 children above 18 years, hence the number of schoolchildren between 6-18 years was 12,228. The percentages for the age groups are calculated based on the 12,228 schoolchildren.
*** Swiss-SEP: Swiss socioeconomic position version 2.0 published by Panczak et al. [10], census data from 2012 - 2015. Reported are the quartiles of Swiss-SEP values observed in the LUIS data and observed in the population of the local growth references [2]. For example, the highest 25% of Swiss-SEP values were >76.5 in LUIS, while the highest 25% of Swiss-SEP values were >72.3 in the population of the local growth references.

**Table S2:**
Mean height, weight, and BMI z-scores based on WHO, local, and IOTF growth references by sex, age, origin of parents, Swiss-SEP, and urbanization; and the fit of z-scores to a standard normal distribution.

| | | | WHO | | | Local | | | IOTF | | |
|---|---|---|---|---|---|---|---|---|---|---|---|
| | | | | Anderson Darling* | | | Anderson Darling* | | | Anderson Darling* | |
| | | n | mean (sd) | $A^2$ | p-value | mean (sd) | $A^2$ | p-value | mean (sd) | $A^2$ | p-value |
| **Sex** | | | | | | | | | | | |
| **Girls** | Height z-score | 1872 | 0.52 (0.98) | 250.1 | <0.001 | 0.15 (1.02) | 23.0 | <0.001 | *NA* | *NA* | *NA* |
| | Weight z-score | 1872 | 0.21 (0.97) | 37.7 | <0.001 | 0.05 (0.99) | 4.2 | 0.007 | *NA* | *NA* | *NA* |
| | BMI z-score | 1872 | 0.02 (1.10) | 5.3 | 0.002 | -0.02 (1.04) | 1.0 | 0.356 | 0.15 (1.05) | 21.6 | <0.001 |
| **Boys** | Height z-score | 1883 | 0.60 (1.02) | 333.7 | <0.001 | 0.16 (1.03) | 25.8 | <0.001 | *NA* | *NA* | *NA* |
| | Weight z-score | 1883 | 0.35 (1.05) | 97.8 | <0.001 | 0.07 (0.98) | 6.7 | <0.001 | *NA* | *NA* | *NA* |
| | BMI z-score | 1883 | 0.10 (1.17) | 23.1 | <0.001 | 0.00 (0.97) | 1.0 | 0.358 | 0.23 (1.04) | 45.0 | <0.001 |
| **Age** | | | | | | | | | | | |
| **6-8** | Height z-score | 678 | 0.82 (0.92) | 232.1 | <0.001 | 0.32 (0.95) | 37.6 | <0.001 | *NA* | *NA* | *NA* |
| | Weight z-score | 678 | 0.44 (0.95) | 62.0 | <0.001 | 0.12 (0.89) | 8.2 | <0.001 | *NA* | *NA* | *NA* |
| | BMI z-score | 678 | -0.04 (1.06) | 2.3 | 0.063 | -0.11 (0.94) | 5.5 | 0.002 | 0.01 (0.97) | 1.0 | 0.368 |
| **9-11** | Height z-score | 881 | 0.51 (1.04) | 106.8 | <0.001 | 0.06 (1.07) | 2.4 | 0.054 | *NA* | *NA* | *NA* |
| | Weight z-score | 881 | 0.12 (0.94) | 6.7 | <0.001 | -0.08 (0.95) | 3.6 | 0.014 | *NA* | *NA* | *NA* |
| | BMI z-score | 881 | -0.05 (1.11) | 5.0 | 0.003 | -0.13 (0.97) | 8.4 | <0.001 | 0.03 (0.98) | 1.5 | 0.182 |
| **12-14** | Height z-score | 1663 | 0.53 (1.01) | 236.2 | <0.001 | 0.19 (1.03) | 31.8 | <0.001 | *NA* | *NA* | *NA* |
| | Weight z-score | 1663 | 0.31 (1.05) | 67.8 | <0.001 | 0.10 (1.02) | 10.8 | <0.001 | *NA* | *NA* | *NA* |
| | BMI z-score | 1663 | 0.13 (1.19) | 28.5 | <0.001 | 0.05 (1.05) | 4.7 | 0.004 | 0.29 (1.09) | 66.7 | <0.001 |
| **15-17** | Height z-score | 533 | 0.39 (0.95) | 41.4 | <0.001 | 0.00 (1.01) | 0.43 | 0.820 | *NA* | *NA* | *NA* |
| | Weight z-score | 533 | 0.24 (1.03) | 13.7 | <0.001 | 0.09 (1.01) | 3.6 | 0.014 | *NA* | *NA* | *NA* |
| | BMI z-score | 533 | 0.18 (1.10) | 8.8 | <0.001 | 0.12 (0.99) | 4.2 | 0.007 | 0.40 (1.04) | 39.6 | <0.001 |
| **Origin of parents**** | | | | | | | | | | | |
| **CH** | Height z-score | 1933 | 0.52 (0.98) | 256.2 | <0.001 | 0.11 (1.00) | 11.9 | <0.001 | *NA* | *NA* | *NA* |
| | Weight z-score | 1933 | 0.14 (0.94) | 18.9 | <0.001 | -0.07 (0.93) | 8.0 | <0.001 | *NA* | *NA* | *NA* |
| | BMI z-score | 1933 | -0.10 (1.06) | 11.7 | <0.001 | -0.15 (0.95) | 23.9 | <0.001 | 0.04 (0.97) | 2.9 | 0.033 |
| **CH1** | Height z-score | 584 | 0.61 (1.02) | 111.9 | <0.001 | 0.19 (1.04) | 13.9 | <0.001 | *NA* | *NA* | *NA* |
| | Weight z-score | 584 | 0.28 (1.04) | 19.8 | <0.001 | 0.04 (1.02) | 1.1 | 0.298 | *NA* | *NA* | *NA* |
| | BMI z-score | 584 | 0.01 (1.16) | 4.7 | 0.004 | -0.06 (1.03) | 1.5 | 0.166 | 0.13 (1.06) | 5.3 | 0.002 |
| **NW** | Height z-score | 203 | 0.77 (1.06) | 58.6 | <0.001 | 0.34 (1.10) | 12.5 | <0.001 | *NA* | *NA* | *NA* |
| | Weight z-score | 203 | 0.27 (0.92) | 7.2 | <0.001 | 0.03 (0.90) | 0.8 | 0.447 | *NA* | *NA* | *NA* |
| | BMI z-score | 203 | -0.12 (1.03) | 2.4 | 0.058 | -0.17 (0.91) | 4.1 | 0.007 | -0.01 (0.94) | 0.8 | 0.453 |
| **SE** | Height z-score | 382 | 0.65 (0.98) | 80.4 | <0.001 | 0.27 (1.00) | 14.2 | <0.001 | *NA* | *NA* | *NA* |
| | Weight z-score | 382 | 0.71 (1.08) | 87.1 | <0.001 | 0.48 (1.00) | 46.9 | <0.001 | *NA* | *NA* | *NA* |
| | BMI z-score | 382 | 0.59 (1.18) | 67.6 | <0.001 | 0.45 (1.02) | 42.9 | <0.001 | 0.70 (1.08) | 91.9 | <0.001 |
| **OM** | Height z-score | 263 | 0.53 (1.14) | 37.8 | <0.001 | 0.15 (1.16) | 5.1 | 0.002 | *NA* | *NA* | *NA* |
| | Weight z-score | 263 | 0.50 (1.06) | 29.9 | <0.001 | 0.28 (1.02) | 12.1 | <0.001 | *NA* | *NA* | *NA* |
| | BMI z-score | 263 | 0.39 (1.23) | 23.7 | <0.001 | 0.28 (1.07) | 12.4 | <0.001 | 0.50 (1.12) | 34.6 | <0.001 |
| **Swiss-SEP groups***** | | | | | | | | | | | |
| **1: 0 - 49** | Height z-score | 138 | 0.49 (0.98) | 17.3 | <0.001 | 0.12 (1.00) | 1.6 | 0.151 | *NA* | *NA* | *NA* |
| | Weight z-score | 138 | 0.55 (1.04) | 19.2 | <0.001 | 0.35 (0.98) | 9.0 | <0.001 | *NA* | *NA* | *NA* |
| | BMI z-score | 138 | 0.49 (1.15) | 16.7 | <0.001 | 0.37 (1.00) | 10.2 | <0.001 | 0.63 (1.05) | 26.6 | <0.001 |
| **2: 40 - 56** | Height z-score | 459 | 0.41 (1.03) | 38.1 | <0.001 | 0.03 (1.05) | 1.1 | 0.292 | *NA* | *NA* | *NA* |
| | Weight z-score | 459 | 0.43 (1.15) | 36.9 | <0.001 | 0.21 (1.11) | 12.6 | <0.001 | *NA* | *NA* | *NA* |
| | BMI z-score | 459 | 0.35 (1.29) | 37.7 | <0.001 | 0.24 (1.14) | 18.8 | <0.001 | 0.49 (1.19) | 58.1 | <0.001 |
| **3: 57 - 62** | Height z-score | 431 | 0.55 (0.92) | 67.1 | <0.001 | 0.15 (0.95) | 5.8 | <0.001 | *NA* | *NA* | *NA* |
| | Weight z-score | 431 | 0.33 (0.99) | 19.8 | <0.001 | 0.11 (0.96) | 3.3 | 0.016 | *NA* | *NA* | *NA* |
| | BMI z-score | 431 | 0.13 (1.15) | 5.3 | 0.002 | 0.05 (1.01) | 1.0 | 0.394 | 0.25 (1.05) | 13.4 | <0.001 |
| **4: 62 - 69** | Height z-score | 841 | 0.52 (1.03) | 108.2 | <0.001 | 0.12 (1.06) | 6.7 | 0.001 | *NA* | *NA* | *NA* |
| | Weight z-score | 841 | 0.23 (1.03) | 18.0 | <0.001 | 0.01 (1.00) | 0.5 | 0.828 | *NA* | *NA* | *NA* |
| | BMI z-score | 841 | 0.03 (1.16) | 5.5 | 0.002 | -0.04 (1.03) | 1.0 | 0.366 | 0.17 (1.06) | 12.0 | <0.001 |
| **5: 70 - 100** | Height z-score | 1886 | 0.62 (1.00) | 359.3 | <0.001 | 0.21 (1.02) | 41.7 | <0.001 | *NA* | *NA* | *NA* |
| | Weight z-score | 1886 | 0.24 (0.96) | 47.4 | <0.001 | 0.02 (0.94) | 3.6 | 0.014 | *NA* | *NA* | *NA* |
| | BMI z-score | 1886 | -0.03 (1.07) | 5.3 | 0.002 | -0.10 (0.94) | 11.5 | <0.001 | 0.09 (0.97) | 7.1 | <0.001 |
| **Urbanization****** | | | | | | | | | | | |
| **Rural** | Height z-score | 247 | 0.39 (1.06) | 19.7 | <0.001 | -0.05 (1.08) | 0.9 | 0.425 | *NA* | *NA* | *NA* |
| | Weight z-score | 247 | -0.09 (0.97) | 2.4 | 0.056 | -0.32 (0.99) | 12.5 | <0.001 | *NA* | *NA* | *NA* |
| | BMI z-score | 247 | -0.38 (1.13) | 18.1 | <0.001 | -0.41 (1.00) | 20.5 | <0.001 | -0.25 (1.02) | 8.6 | <0.001 |
| **Small urban** | Height z-score | 1635 | 0.54 (0.99) | 230.6 | <0.001 | 0.16 (1.02) | 21.1 | <0.001 | *NA* | *NA* | *NA* |
| | Weight z-score | 1635 | 0.26 (1.02) | 44.0 | <0.001 | 0.05 (0.99) | 2.8 | 0.036 | *NA* | *NA* | *NA* |
| | BMI z-score | 1635 | 0.06 (1.13) | 10.5 | <0.001 | -0.01 (1.00) | 0.3 | 0.901 | 0.21 (1.04) | 33.7 | <0.001 |
| **Large urban** | Height z-score | 1873 | 0.60 (1.00) | 334.3 | <0.001 | 0.18 (1.02) | 32.6 | <0.001 | *NA* | *NA* | *NA* |
| | Weight z-score | 1873 | 0.35 (0.99) | 100.7 | <0.001 | 0.12 (0.96) | 17.6 | <0.001 | *NA* | *NA* | *NA* |
| | BMI z-score | 1873 | 0.13 (1.13) | 20.4 | <0.001 | 0.04 (1.00) | 2.5 | <0.001 | 0.23 (1.04) | 47.4 | <0.001 |

* Anderson-Darling test to assess goodness of fit of the observed z-score distribution based on the WHO, local, or IOTF growth references to a standard normal distribution, $A^2$: test statistic for goodness of fit, Color coding: green: best fit in respective group.
** Origin of parents: CH: both parents from Switzerland; CH1: one parent from Switzerland; NW: both parents from Northern/Western Europe; SE: both parents from Southern/Eastern Europe; OM: Other/Mixed, parents from other or not from the same regions. For the origin of parents, we included n = 3365 children.
*** Swiss-SEP: Swiss socioeconomic position version 2.0, census data from 2012 - 2015. The 5 groups are based on quintiles published by Panczak et al. [10]. A higher Swiss-SEP relates to a higher socioeconomic position.
**** Urbanization: rural: rural areas; small urban: towns and suburbs; large urban: cities.
BMI: Body mass index; IOTF: International Obesity Task Force; NA: not applicable; sd: standard deviation; WHO: World Health Organization.

**Table S3:**
Linear regression of height z-scores based on local references on age, sex, origin of parents, Swiss-SEP, and urbanization.

| | Height z-score | | | | | | | |
|---|---|---|---|---|---|---|---|---|
| | Univariable model | | | | Multivariable model | | | |
| | **β-coefficient (95% CI)** | **std. error** | **p-value** | **adj. $R^2$** | **β-coefficient (95% CI)** | **std. error** | **p-value** | **adj. $R^2$** |
| **Age** | | | | | | | | |
| (1 year increase) | -0.03 (-0.04, -0.01) | 0.01 | <0.001 | 0.005 | -0.03 (-0.05, -0.02) | 0.008 | <0.001 | |
| **Sex** | | | | | | | | |
| Girls | ref | ref | ref | | *NA* | *NA* | *NA* | |
| Boys | 0.01 (-0.06, 0.07) | 0.03 | 0.815 | 0 | *NA* | *NA* | *NA* | |
| **Origin of parents*** | | | | | | | | |
| Switzerland (both parents) | ref | ref | ref | | ref | ref | ref | |
| Switzerland (one parent) | 0.08 (-0.01, 0.18) | 0.05 | 0.086 | | 0.06 (-0.03, 0.16) | 0.05 | 0.188 | |
| Northern/Western Europe | 0.23 (0.08, 0.38) | 0.08 | 0.003 | | 0.19 (0.04, 0.33) | 0.08 | 0.015 | 0.013 |
| Southern/Eastern Europe | 0.16 (0.04, 0.27) | 0.06 | 0.006 | | 0.22 (0.10, 0.34) | 0.06 | <0.001 | |
| Other/Mixed | 0.04 (-0.10, 0.17) | 0.07 | 0.585 | 0.003 | 0.08 (-0.06, 0.21) | 0.07 | 0.263 | |
| **Swiss-SEP**** | | | | | | | | |
| (per increment of 20) | 0.10 (0.04, 0.16) | 0.04 | <0.001 | 0.003 | 0.06 (-0.02, 0.14) | 0.04 | 0.125 | |
| **Urbanization***** | | | | | | | | |
| Rural | ref | ref | ref | | ref | ref | ref | |
| Small urban | 0.21 (0.07, 0.35) | 0.07 | 0.003 | | 0.27 (0.12, 0.42) | 0.08 | <0.001 | |
| Large urban | 0.23 (0.09, 0.37) | 0.07 | 0.001 | 0.002 | 0.20 (0.05, 0.35) | 0.08 | 0.010 | |

* Origin of parents: For the univariable model on origin of parents and for the multivariable model we included n = 3365 children.
** Swiss-SEP: Swiss socioeconomic position version 2.0, census data from 2012 - 2015. A higher score relates to a higher socioeconomic position.
*** Urbanization: rural: rural areas; small urban: towns and suburbs; large urban: cities.
adj: adjusted; NA: not applicable, not included in multivariable model based on results of univariable model. ref: reference.

**Table S4:**
Linear regression of weight z-scores based on local references on age, sex, origin of parents, Swiss-SEP, and urbanization.

| | Weight z-score | | | | | | | |
|---|---|---|---|---|---|---|---|---|
| | Univariable model | | | | Multivariable model | | | |
| | **β-coefficient (95% CI)** | **std. error** | **p-value** | **adj. $R^2$** | **β-coefficient (95% CI)** | **std. error** | **p-value** | **adj. $R^2$** |
| **Age** | | | | | | | | |
| (1 year increase) | 0.01 (-0.00, 0.02) | 0.01 | 0.221 | 0 | *NA* | *NA* | *NA* | |
| **Sex** | | | | | | | | |
| Female | ref | ref | ref | | *NA* | *NA* | *NA* | |
| Male | 0.01 (-0.05, 0.07) | 0.03 | 0.732 | 0 | *NA* | *NA* | *NA* | |
| **Origin of parents*** | | | | | | | | |
| Switzerland (both parents) | ref | ref | ref | | ref | ref | ref | |
| Switzerland (one parent) | 0.12 (0.03, 0.21) | 0.05 | 0.013 | | 0.10 (0.01, 0.19) | 0.05 | 0.034 | |
| Northern/Western Europe | 0.10 (-0.03, 0.24) | 0.07 | 0.141 | | 0.10 (-0.04, 0.24) | 0.07 | 0.167 | 0.044 |
| Southern/Eastern Europe | 0.55 (0.45, 0.66) | 0.05 | <0.01 | | 0.49 (0.38, 0.60) | 0.06 | <0.001 | |
| Other/Mixed | 0.36 (0.23, 0.48) | 0.06 | <0.01 | 0.034 | 0.30 (0.17, 0.43) | 0.06 | <0.001 | |
| **Swiss-SEP**** | | | | | | | | |
| (per increment of 20) | -0.12 (-0.18, -0.007) | 0.03 | <0.001 | 0.004 | -0.08 (-0.15, -0.01) | 0.04 | 0.019 | |
| **Urbanization***** | | | | | | | | |
| Rural | ref | ref | ref | | ref | ref | ref | |
| Small urban | 0.37 (0.24, 0.50) | 0.07 | <0.001 | | 0.39 (0.25, 0.53) | 0.07 | <0.001 | |
| Large urban | 0.45 (0.32, 0.58) | 0.07 | <0.001 | 0.012 | 0.42 (0.27, 0.56) | 0.07 | <0.001 | |

* Origin of parents: For the univariable model on origin of parents and for the multivariable model we included n = 3365 children.
** Swiss-SEP: Swiss socioeconomic position version 2.0, census data from 2012 - 2015. A higher score relates to a higher socioeconomic position.
*** Urbanization: rural: rural areas; small urban: towns and suburbs; large urban: cities.
adj: adjusted; NA: not applicable, not included in multivariable model based on results of univariable model. ref: reference.

**Table S5:**
Linear regression of BMI z-scores based on local references on age, sex, origin of parents, Swiss-SEP, and urbanization.

| BMI z-score | | | | | | | | |
|---|---|---|---|---|---|---|---|---|
| | **Univariable model** | | | | **Multivariable model** | | | |
| | **β-coefficient (95% CI)** | **std. error** | **p-value** | **adj. $R^2$** | **β-coefficient (95% CI)** | **std. error** | **p-value** | **adj. $R^2$** |
| **Age** | | | | | | | | |
| (1 year increase) | 0.04 (0.02, 0.05) | 0.01 | <0.001 | 0.009 | 0.01 (-0.01, 0.02) | 0.01 | 0.342 | |
| **Sex** | | | | | | | | |
| Female | ref | ref | ref | | *NA* | *NA* | *NA* | |
| Male | 0.01 (-0.05, 0.08) | 0.03 | 0.702 | 0 | *NA* | *NA* | *NA* | |
| **Origin of parents*** | | | | | | | | |
| Switzerland (both parents) | ref | ref | ref | | ref | ref | ref | |
| Switzerland (one parent) | 0.09 (0.00, 0.18) | 0.05 | 0.046 | | 0.07 (-0.02, 0.17) | 0.05 | 0.106 | |
| Northern/Western Europe | -0.02 (-0.16, 0.12) | 0.07 | 0.805 | | -0.00 (-0.15, 0.14) | 0.07 | 0.945 | 0.057 |
| Southern/Eastern Europe | 0.61 (0.50, 0.72) | 0.05 | <0.001 | | 0.49 (0.38, 0.61) | 0.06 | <0.001 | |
| Other/Mixed | 0.43 (0.31, 0.56) | 0.06 | <0.001 | 0.043 | 0.34 (0.22, 0.47) | 0.07 | <0.001 | |
| **Swiss-SEP**** | | | | | | | | |
| (per increment of 20) | -0.22 (-0.28, -0.16) | 0.03 | <0.001 | 0.004 | -0.16 (-0.24, -0.09) | 0.04 | <0.001 | |
| **Urbanization***** | | | | | | | | |
| Rural | ref | ref | ref | | ref | ref | ref | |
| Small urban | 0.40 (0.27, 0.53) | 0.07 | <0.001 | | 0.43 (0.29, 0.58) | 0.07 | <0.001 | |
| Large urban | 0.45 (0.32, 0.59) | 0.07 | <0.001 | 0.012 | 0.46 (0.31, 0.60) | 0.07 | <0.001 | |

* Origin of parents: For the univariable model on origin of parents and for the multivariable model we included n = 3365 children.
** Swiss-SEP: Swiss socioeconomic position version 2.0, census data from 2012 - 2015. A higher score relates to a higher socioeconomic position.
*** Urbanization: rural: rural areas; small urban: towns and suburbs; large urban: cities.
adj: adjusted; BMI: body mass index; NA: not applicable, not included in multivariable model based on results of univariable model. ref: reference.

**Table S6.** Cutoff values for the BMI and height categories based on WHO, local, and IOTF growth references. Number of children and proportions in BMI and height categories in 3755 children of the LuftiBus in the school (LUIS) study.

| BMI category | WHO | | | Local | | | | IOTF | | | |
|---|---|---|---|---|---|---|---|---|---|---|---|
| | cutoff* | n | % (95% CI) | cutoff* | | n | % (95% CI) | cutoff* | | n | % (95% CI) |
| | | | | Girls | Boys | | | Girls | Boys | | |
| **Severe obesity** | >99.5 | 62 | 1.7 (1.3, 2.1) | >99.3 | >98.8 | 22 | 0.6 (0.4, 0.9) | >99.8 | >99.8 | 11 | 0.3 (0.2, 0.5) |
| **Obesity** | >97 | 171 | 4.6 (3.9, 5.3) | >96.8 | >95.5 | 131 | 3.5 (2.9, 4.1) | >98.6 | >98.9 | 94 | 2.5 (2.0, 3.1) |
| **Overweight** | >90 | 318 | 8.5 (7.6, 9.4) | >82.9 | >78.9 | 568 | 15.1 (14.0, 16.3) | >89.3 | >90.5 | 472 | 12.6 (11.5, 13.7) |
| **Normal weight** | ≤90 | 3204 | 85.3 (84.1, 86.4) | ≤82.9 | ≤78.9 | 3034 | 80.8 (79.5, 82.0) | ≤89.3 | ≤90.5 | 3178 | 84.6 (83.4, 85.8) |
| **Cohen's Kappa (95%CI)**** | ref | | | 0.74 (0.71, 0.76) | | | | 0.73 (0.71, 0.76) | | | |

| Height category | WHO | | | Local | | | IOTF |
|---|---|---|---|---|---|---|---|
| | cutoff* | n | % (95% CI) | cutoff* | n | % (95% CI) | |
| **Tall stature** | >97 | 330 | 8.8 (7.9, 9.8) | >97 | 330 | 4.3 (3.7, 5.0) | *NA* |
| **Normal** | ≤≥ | 3372 | 90.1 (89.1, 91.0) | ≤≥ | 3372 | 92.8 (91.9, 93.6) | |
| **Short stature** | <3 | 41 | 1.1 (0.8, 1.5) | <3 | 41 | 2.9 (2.4, 3.5) | |
| **Cohen's Kappa (95%CI)**** | ref | | | 0.60 (0.56, 0.65) | | | |

* Cutoffs for BMI and height categories are given in percentiles. We used the cutoffs published by WHO, local, and IOTF growth references to define BMI categories.
** Cohen's Kappa to quantify agreement of BMI and height categories between WHO compared to local or compared to IOTF growth references. We interpreted Cohen's Kappa between 0.61-0.80 as substantial and 0.41-0.60 as moderate agreement, according to Landis and Koch [11].
BMI: Body mass index; IOTF: International Obesity Task Force; NA: not applicable; ref: reference; WHO: World Health Organization.

**Table S7.** Agreement of BMI and height classification* by WHO compared to local and IOTF growth references by sex, age, origin of parents, and Swiss-SEP.

| | | n | WHO Cohen's Kappa** (95% CI) | Local Cohen's Kappa** (95% CI) | IOTF Cohen's Kappa** (95% CI) |
|---|---|---|---|---|---|
| **Sex** | | | | | |
| **Female** | BMI category | 1872 | ref | 0.74 (0.70, 0.78) | 0.76 (0.72, 0.80) |
| | Height category | 1872 | ref | 0.64 (0.57, 0.70) | *NA* |
| **Male** | BMI category | 1883 | ref | 0.74 (0.70, 0.77) | 0.71 (0.67, 0.74) |
| | Height category | 1883 | ref | 0.57 (0.51, 0.63) | *NA* |
| **Age (years)** | | | | | |
| **6-8** | BMI category | 678 | ref | 0.70 (0.63, 0.77) | 0.72 (0.65, 0.79) |
| | Height category | 678 | ref | 0.53 (0.42, 0.64) | *NA* |
| **9-11** | BMI category | 881 | ref | 0.76 (0.71, 0.82) | 0.63 (0.57, 0.70) |
| | Height category | 881 | ref | 0.57 (0.48, 0.66) | *NA* |
| **12-14** | BMI category | 1663 | ref | 0.75 (0.71, 0.78) | 0.77 (0.74, 0.80) |
| | Height category | 1663 | ref | 0.66 (0.60, 0.73) | *NA* |
| **15-17** | BMI category | 533 | ref | 0.72 (0.65, 0.78) | 0.71 (0.65, 0.78) |
| | Height category | 533 | ref | 0.56 (0.42, 0.70) | *NA* |
| **Origin of parents*** ** | | | | | |
| **Switzerland (both parents)** | BMI category | 1933 | ref | 0.72 (0.68, 0.76) | 0.74 (0.70, 0.78) |
| | Height category | 1933 | ref | 0.60 (0.53, 0.67) | *NA* |
| **Switzerland (one parent)** | BMI category | 584 | ref | 0.79 (0.73, 0.85) | 0.71 (0.64, 0.77) |
| | Height category | 584 | ref | 0.53 (0.41, 0.65) | *NA* |
| **Northern/Western Europe** | BMI category | 203 | ref | 0.69 (0.54, 0.84) | 0.72 (0.58, 0.86) |
| | Height category | 203 | ref | 0.74 (0.60, 0.88) | *NA* |
| **Southern/Eastern Europe** | BMI category | 382 | ref | 0.73 (0.67, 0.79) | 0.70 (0.64, 0.77) |
| | Height category | 382 | ref | 0.67 (0.54, 0.80) | *NA* |
| **Other/ Mixed** | BMI category | 263 | ref | 0.74 (0.66, 0.81) | 0.76 (0.68, 0.83) |
| | Height category | 263 | ref | 0.61 (0.46, 0.76) | *NA* |
| **Swiss-SEP 2.0 quintiles******** | | | | | |
| **1: 0 - 49** | BMI category | 138 | ref | 0.71 (0.60, 0.82) | 0.74 (0.63, 0.85) |
| | Height category | 138 | ref | 0.64 (0.40, 0.89) | *NA* |
| **2: 40 - 56** | BMI category | 459 | ref | 0.73 (0.67, 0.79) | 0.70 (0.64, 0.76) |
| | Height category | 459 | ref | 0.54 (0.40, 0.67) | *NA* |
| **3: 57 - 62** | BMI category | 427 | ref | 0.78 (0.71, 0.85) | 0.73 (0.66, 0.81) |
| | Height category | 427 | ref | 0.45 (0.29, 0.61) | *NA* |
| **4: 62 - 69** | BMI category | 859 | ref | 0.75 (0.70, 0.80) | 0.75 (0.69, 0.80) |
| | Height category | 859 | ref | 0.64 (0.55, 0.73) | *NA* |
| **5: 70 - 100** | BMI category | 1872 | ref | 0.72 (0.68, 0.76) | 0.73 (0.68, 0.77) |
| | Height category | 1872 | ref | 0.62 (0.56, 0.69) | *NA* |
| **Urbanization******* | | | | | |
| **Rural** | BMI category | 247 | ref | 0.76 (0.62, 0.90) | 0.75 (0.62, 0.89) |
| | Height category | 247 | ref | 0.57 (0.41, 0.73) | *NA* |
| **Small urban** | BMI category | 1635 | ref | 0.75 (0.71, 0.79) | 0.75 (0.72, 0.79) |
| | Height category | 1635 | ref | 0.62 (0.55, 0.69) | *NA* |
| **Large urban** | BMI category | 1873 | ref | 0.73 (0.69, 0.76) | 0.71 (0.67, 0.74) |
| | Height category | 1873 | ref | 0.59 (0.53, 0.66) | *NA* |

* BMI and height categories: We classified BMI and height into categories as shown in Table S6.
** Cohen's Kappa to quantify agreement of BMI and height categories between WHO compared to local or IOTF growth references. We interpreted Cohen's Kappa between 0.61-0.80 as substantial (color: green) and 0.41-0.60 as moderate (color: red) agreement, according to Landis and Koch [11].
*** For the origin of parents, we included n = 3365 children.
**** Swiss-SEP: Swiss socioeconomic position version 2.0, census data from 2012 - 2015. The 5 groups are based on quintiles published by Panczak et al. [10]. A higher score relates to a higher socioeconomic position.
***** Urbanization: rural: rural areas; small urban: towns and suburbs; large urban: cities.
BMI: Body mass index; IOTF: International Obesity Task Force; NA: not applicable; ref: reference; WHO: World Health Organization.
